# Coverage and Systemic Barriers to Maternal Micronutrient Supplementation in Bayelsa State, Nigeria

**DOI:** 10.1101/2025.07.15.25331588

**Authors:** Mordecai Oweibia, Tarimobowei Egberipou, Gift Cornelius Timighe, Ebiakpor Bainkpo Agbedi, Zuofa Seimo Egberipou, Ekadi Francis Tamaradielaye

**Author notes:** Corresponding Author Mordecai Oweibia, *FIMC, FRSPH. FAIPH.* Department of Public Health, Bayelsa Medical University, Yenagoa, Nigeria. |.

## Abstract

**Introduction:** Maternal micronutrient supplementation (MMS) is a globally endorsed strategy to combat nutritional deficiencies during pregnancy and improve birth outcomes. Despite strong evidence supporting its integration into antenatal care services, coverage in many low-resource settings remains suboptimal. This study assesses the coverage and systemic challenges associated with MMS delivery during the June 2025 Maternal and Child Health (MNCH) Week in Bayelsa State, Nigeria.

**Methodology:** A descriptive cross-sectional design was employed, using validated secondary data from the OPS Room Final Report and Power BI dashboards covering all eight LGAs in Bayelsa State. Analytical methods included percentage-based coverage assessment, comparative ratio analysis with other interventions (Vitamin A and Deworming), and evaluation of implementation gaps using discrepancy indices. Operational notes from State Technical Facilitators were triangulated to contextualize the quantitative findings.

**Results:** Out of an estimated 6,982 pregnant women, only 2,197 received either MMS or Iron-Folate supplementation, resulting in a coverage rate of 31.5%. This figure was significantly lower than coverage for Vitamin A (83%) and Deworming (55%). Operational challenges identified included delayed rollout in Southern Ijaw, incomplete data reporting in Nembe and SILGA, non-disaggregated documentation between MMS and IFA, and limited awareness among beneficiaries. These systemic weaknesses significantly constrained the delivery and visibility of maternal nutrition services during the campaign.

**Conclusion:** The findings reveal that despite the large-scale nature of MNCH Week, maternal supplementation remains poorly prioritized and inconsistently delivered. Without targeted reforms in training, documentation, logistics, and digital monitoring, MMS will continue to underperform compared to child-focused services. Improving maternal outcomes requires elevating MMS within campaign structures and routine antenatal care systems to ensure equitable and effective service reach.

## 1.0 INTRODUCTION

Micronutrient deficiencies remain a major public health challenge in low- and middle-income countries (LMICs), including Nigeria, where vulnerable populations such as pregnant women are disproportionately affected due to increased nutritional demands and limited dietary diversity (Global Nutrition Report, 2021; National Bureau of Statistics & UNICEF, 2021). The physiological demands of pregnancy require elevated intake of specific micronutrients such as iron, folate, iodine, vitamin A, and zinc to support maternal health and fetal development and to prevent complications like anemia, preeclampsia, low birth weight, and neural tube defects (Black *et al.,* 2013; WHO, 2020).

Despite longstanding antenatal care (ANC) programs, deficiencies in these key micronutrients remain widespread in Nigeria. The Nigeria Multiple Indicator Cluster Survey (MICS) 2021 revealed alarming rates of iron and folate deficiencies, particularly in the South-South and North-East regions (National Bureau of Statistics & UNICEF, 2021). To address this, the World Health Organization (WHO) endorses Multiple Micronutrient Supplementation (MMS) as a superior alternative to Iron-Folic Acid (IFA) alone, especially in LMICs with high maternal malnutrition and anemia rates. Meta-analyses have demonstrated that MMS significantly reduces the risk of low birth weight, small-for-gestational-age births, and neonatal mortality when compared to IFA-only interventions (Keats *et al.,* 2019; Smith *et al.,* 2017; WHO, 2020).

However, in Nigeria, MMS adoption remains fragmented and inconsistent across states. A recent review by the Nigerian Federal Ministry of Health (FMOH) highlights that only a limited number of states have piloted MMS in routine ANC or campaign-based service delivery, and national rollout guidelines are still under development (FMOH, 2023). This is concerning given that anemia prevalence among pregnant women remains persistently high, with 61% of pregnant Nigerian women found to be anemic, according to the Nigeria Demographic and Health Survey (NDHS, 2018). The continued reliance on IFA-only supplementation, despite such statistics, suggests that implementation of comprehensive nutrition interventions is falling short of addressing the broader spectrum of micronutrient needs (Oweibia *et al.,* 2025).

In Bayelsa State, known for its challenging riverine terrain and difficult health logistics, MMS implementation faces further complications. Many local government areas (LGAs) are only accessible by boat, and health workers report recurring delays in logistics, stock deployment, and service delivery (Journal of Public Health in Africa, 2022; Bayelsa State Primary Health Care Board Report, 2023). These geographical barriers are compounded by gaps in data management and inconsistent integration of MMS into routine ANC platforms. According to field summaries and dashboards from the OPS Room during the June 2025 Maternal and Child Health (MNCH) Week, MMS was one of the least documented interventions compared to child-focused services like Vitamin A supplementation and Deworming (NPHCDA, 2025).

Globally, MMS has gained traction not only due to its clinical benefits but also its economic advantages. A microsimulation study by Horton et al. (2018) found that MMS yields higher cost-effectiveness and economic return than IFA alone when scaled across LMIC populations (Horton *et al.,* 2018). Additional evidence supports these findings (Young *et al.,* 2022). Nonetheless, full implementation is often hindered by systemic challenges including fragmented supply chains, inconsistent ANC attendance, inadequate pre-campaign training, and weak community demand (Bhutta *et al.,* 2018; WHO, 2020). These challenges are further intensified in humanitarian and remote contexts, where health infrastructure is minimal and program planning is frequently reactive (UNICEF, 2022; UNHCR, 2021).

The case for MMS in Nigeria is further supported by international evidence from countries like Bangladesh, Nepal, and Vietnam, where national adoption of MMS led to improved maternal and neonatal outcomes (Rana & Clift, 2023; Fernández-Gaxiola *et al.,* 2024). In these countries, clear policy commitment, early stakeholder buy-in, and robust supply chain systems contributed to MMS coverage rates exceeding 70%. The use of standardized UNIMMAP-based MMS formulations, as promoted by global nutrition alliances, has provided a scalable template for national adaptation (Ajello *et al.,* 2022).

Nevertheless, shifting from IFA to MMS requires more than a product substitution; it demands comprehensive health system strengthening. Policy harmonization, sustained funding, community engagement, workforce capacity-building, and digital tracking systems must work in concert to achieve meaningful impact (Kruk et al., 2015; King et al., 2020). Without these systemic reforms, coverage will remain low and the opportunity to address maternal nutrition inequities will be lost. Given Nigeria’s status among countries with the highest maternal and neonatal mortality rates globally, this is an urgent public health investment that cannot be deferred (World Bank, 2020; FMOH, 2023).

Accordingly, this study investigates the coverage and implementation barriers of MMS during the June 2025 MNCH Week in Bayelsa State. Using validated OPS Room data, it assesses the actual reach of the intervention, compares it with other services, and highlights operational bottlenecks that undermine its delivery. The findings aim to inform national strategies for transitioning from campaign-based to routine, system-integrated antenatal MMS distribution.

### 1.2 Statement of the Problem

Maternal nutrition remains an under-addressed priority within Nigeria’s public health system, despite its well-documented link to maternal and neonatal survival outcomes. Although antenatal care (ANC) services routinely include Iron-Folic Acid (IFA) supplementation, the actual coverage, adherence, and quality of delivery remain inconsistent across states and regions (NDHS, 2018; Oweibia *et al.,* 2025; FMOH, 2023). National data reveal that 61% of pregnant Nigerian women are anemic, underscoring the inadequacy of current interventions in addressing the broader micronutrient needs during pregnancy (NDHS, 2018; NBS & UNICEF, 2021).

The June 2025 Maternal and Child Health (MNCH) Week campaign in Bayelsa State reported the distribution of 2,197 Iron-Folate supplements to pregnant women. However, there was no disaggregated reporting to identify how many received the full Multiple Micronutrient Supplementation (MMS) formulation (NPHCDA, 2025). This lack of clarity represents a significant data gap that impairs the ability to evaluate performance, monitor equity in service delivery, or inform evidence-based decision-making at the subnational level.

Field reports from the campaign revealed major inconsistencies in MMS rollout across local government areas (LGAs), with some communities receiving no supplies at all, while others lacked trained personnel capable of administering or documenting the intervention correctly (Bayelsa PHC Report, 2023; UNICEF, 2022). These inconsistencies point to systemic failures in supply chain management, health worker capacity-building, and pre-campaign planning, factors that are essential for the successful integration of MMS into public health programming (Bhutta *et al.,* 2018; Gomes *et al.,* 2020).

Moreover, the relatively high uptake of IFA despite persistent maternal anemia raises questions about policy coherence and implementation quality. The continued underperformance of MMS, in contrast, suggests deeper institutional issues such as inadequate financing, poor coordination between ANC and outreach services, and limited awareness among beneficiaries. Cultural and gender-related barriers, including low maternal autonomy and limited male involvement in maternal health decisions, may also affect demand and utilization of new interventions (UNFPA, 2021; Kruk *et al.,* 2015).

Without transparent, disaggregated, and routine monitoring of MMS coverage, national progress toward maternal nutrition goals will remain poorly measured and misrepresented. This documentation gap risks perpetuating a cycle in which interventions appear nominally implemented but remain ineffective at population scale. Unless urgently addressed through system-level reforms and robust accountability mechanisms, these gaps will continue to undermine maternal health outcomes and perpetuate preventable adverse birth events.

### 1.3 Aim and Objectives of the Study

#### Aim

To assess the coverage and implementation challenges of maternal micronutrient supplementation (MMS) among pregnant women during the June 2025 Maternal and Child Health (MNCH) Week in Bayelsa State, Nigeria.

#### Objectives

1. To determine the MMS coverage rates among pregnant women in Bayelsa State during the June 2025 MNCH Week.
2. To explore factors contributing to the relatively high uptake of Iron-Folate supplements, despite persistent anemia prevalence.
3. To identify systemic and operational challenges affecting the effective delivery, documentation, and sustainability of MMS interventions.

### 1.4 Research Questions

1. What is the actual coverage of MMS among pregnant women during the June 2025 MNCH Week in Bayelsa State?
2. What factors explain the relatively high uptake of Iron-Folate supplements despite existing challenges?
3. What systemic and operational barriers hinder effective delivery and documentation of MMS during health campaigns?

### 1.5 Justification of the Study

This study is justified by the need to critically examine the delivery and systems-level integrity of maternal nutrition programs in Nigeria, particularly as the country prepares to transition from Iron-Folic Acid (IFA) supplementation to the WHO-endorsed Multiple Micronutrient Supplementation (MMS) model (WHO, 2020). While MMS has been shown to reduce adverse birth outcomes and improve maternal health in several low- and middle-income countries (Smith *et al.,* 2017; Bhutta et al., 2018), implementation in Nigeria remains fragmented and under-documented (FMOH, 2023). By analyzing validated OPS Room data from the June 2025 MNCH Week in Bayelsa State, this study provides context-specific evidence to guide national scale-up efforts. It also addresses critical data gaps around coverage and operational barriers, which have constrained the effectiveness of antenatal supplementation services (Gomes *et al.,* 2020). The research aligns with national policy goals outlined in Nigeria’s maternal nutrition strategy (FMOH, 2023) and contributes toward achieving Sustainable Development Goals 2 and 3 by highlighting the systemic inequities that affect access to nutrition services among pregnant women in underserved communities (Global Nutrition Report, 2021; UNICEF, 2022).

### 1.6 Significance of the Study

This study contributes critical evidence to the discourse on maternal health and antenatal nutrition in Nigeria by uncovering gaps in MMS implementation and reporting using validated program data. It highlights potential entry points for improving antenatal care (ANC) service quality, particularly through enhanced logistics coordination and targeted training of frontline health workers. Moreover, the study supports both national and global advocacy for transitioning from Iron-Folic Acid (IFA) supplementation to Multiple Micronutrient Supplementation (MMS), by presenting context-specific implementation challenges and success indicators. The findings also provide a foundation for informing health policy reforms aimed at integrating maternal nutrition interventions into both routine ANC services and campaign-based delivery platforms like the MNCH Week.

### 1.7 Scope of the Study

The scope of this study is limited to Bayelsa State, Nigeria, and focuses solely on maternal micronutrient supplementation (MMS) during the June 2025 round of the MNCH Week. The study examines MMS coverage, compares it to Iron-Folate uptake, and explores operational constraints in MMS implementation. It does not include a clinical assessment of supplement efficacy or long-term maternal outcomes. Instead, it uses programmatic and administrative data to evaluate coverage, delivery mechanisms, and reporting integrity. The study also provides comparative insights with other health interventions conducted during the same campaign week.

### 1.8

**Table 1:** Definition of Terms.

| Term | Definition |
| --- | --- |
| MMS (Multiple Micronutrient Supplementation) | A standardized supplement containing 15 essential vitamins and minerals recommended for pregnant women to prevent multiple deficiencies. |
| IFA (Iron-Folic Acid) | A combination supplement of iron and folic acid used to prevent maternal anemia and neural tube defects during pregnancy. |
| MNCH Week | A biannual nationwide health campaign in Nigeria offering maternal and child health interventions, including vaccinations and nutrition. |
| OPS Room | An operational intelligence unit that tracks service delivery and performance metrics in real-time during campaigns. |
| Coverage Rate | The proportion of the target population that received a specific health intervention during a campaign period. |
| ANC (Antenatal Care) | Healthcare services provided to pregnant women to ensure a healthy pregnancy and reduce complications. |
| Red MUAC | A classification in Mid-Upper Arm Circumference screening indicating severe acute malnutrition requiring urgent intervention. |
| STF (State Technical Facilitators) | Health professionals responsible for monitoring, supervising, and validating service delivery during MNCH Week campaigns. |
| NHMIS (National Health Management Information System) | Nigeria’s official data system for health statistics reporting. |
| DHIS2 | A web-based health information system used globally to manage and visualize health data. |
| Disaggregated Data | Data that is broken down by categories such as age, sex, or location to enable targeted analysis and intervention. |
| Supply Chain Gaps | Disruptions or inefficiencies in the distribution of health commodities, leading to stockouts or delivery delays. |

## 2.0 METHODOLOGY

### 2.1 Study Design

This study adopted a descriptive cross-sectional design, using secondary data from the June 2025 Maternal and Child Health (MNCH) Week in Bayelsa State, Nigeria. This design enabled a snapshot assessment of maternal micronutrient supplementation (MMS) performance across all participating Local Government Areas (LGAs). It was chosen for its utility in evaluating program coverage, identifying delivery gaps, and analyzing implementation discrepancies in a time-bound intervention. The study further integrated mixed-method triangulation by combining quantitative data from validated dashboards with operational narratives from State Technical Facilitators (STFs), thereby enhancing internal validity (Kruk *et al.,* 2015).

### 2.2 Study Area

Bayelsa State, located in Nigeria’s South-South geopolitical zone, comprises eight LGAs: Brass, Ekeremor, Kolokuma/Opokuma, Nembe, Ogbia, Sagbama, Southern Ijaw, and Yenagoa. The state is predominantly riverine, with over 60% of communities accessible only by water transport, which presents recurring challenges to health service delivery and logistics (Bayelsa PHC Report, 2023). These access barriers have been widely documented as key constraints in health system resilience in LMICs and significantly affect service coverage, especially during short-term outreach campaigns (UNICEF, 2022; Kruk *et al.,* 2015).

### 2.3 Study Population

The target population included all pregnant women who were eligible to receive either Multiple Micronutrient Supplementation (MMS) or Iron-Folic Acid (IFA) supplementation during the June 2025 MNCH Week in Bayelsa State. For comparative insight, the study also referenced program data for children aged 6–59 months who received Vitamin A supplementation and Deworming tablets during the same campaign period.

The estimated total number of pregnant women was 6,982, derived using the World Health Organization’s pregnancy estimation factor of 4.5% of the total population. This factor has been widely used in reproductive health programming to estimate the number of expected pregnancies within a given population (WHO, 2022; UNAIDS, 2011). This initial estimate was further adjusted using community-level headcounts, LGA-specific antenatal attendance data, and routine ANC registers validated by the Bayelsa State Primary Health Care Board. These adjustments ensured contextual accuracy and alignment with recent demographic shifts and service utilization trends.

### 2.4 Data Source and Collection

Data for this study were obtained from the OPS Room Final Report and Power BI digital dashboards maintained by the Bayelsa State MNCH Week coordination team. Primary data entry was carried out at the ward level by trained State Technical Facilitators (STFs) and facility-based health workers, using standardized paper-based tally sheets. These were subsequently verified and uploaded to the Power BI platform through structured data flows.

A data reconciliation process was instituted to ensure internal consistency and minimize reporting errors. Discrepancies between the paper-based tallies and the digital entries were flagged during initial data reviews, escalated to LGA Monitoring & Evaluation (M&E) teams for verification, and resolved in alignment with national data quality assurance protocols. Final entries were validated and locked by 10:27 AM on June 23, 2025. This process followed guidance from the WHO Data Quality Review Toolkit (WHO, 2017) and the DHIS2 Implementation Handbook on facility-level data integrity (HISP/WHO, 2019).

The following variables were extracted for analysis:

- Number of pregnant women supplemented with MMS and/or IFA
- Estimated ANC-eligible population per LGA
- Ward-level supplement distribution breakdowns
- Documentation of delayed rollout, tool deployment gaps, and STF reporting inconsistencies
- Coverage data for Vitamin A and Deworming interventions as comparator indicators

All data were anonymized and aggregated at the LGA level, ensuring full compliance with ethical standards for secondary data analysis and digital health data protection.

### 2.5 Analytical Framework

The analysis employed three main approaches: (1) coverage rate estimation, (2) comparative ratio analysis, and (3) discrepancy index computation, as described below:

#### 2.5.1 Coverage Rate Calculation

Coverage rates were calculated using the standard epidemiological formula:

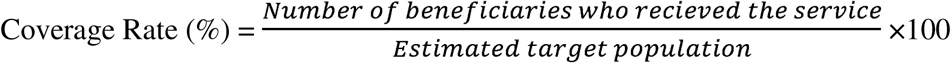

This formula was applied to MMS/IFA, Vitamin A, and Deworming data to determine uptake proportions across service categories.

#### 2.5.2 Comparative Uptake Ratio

To assess the relative performance of MMS against other interventions, the following ratio was used:

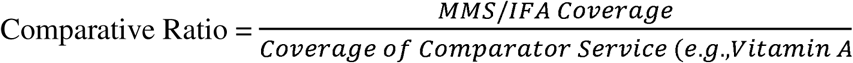

A value <1 indicates underperformance compared to the reference service.

#### 2.5.3 Discrepancy Index for Systemic Gaps

To identify inconsistencies in implementation and reporting, a modified WHO Data Quality Audit (DQA) discrepancy index was used:

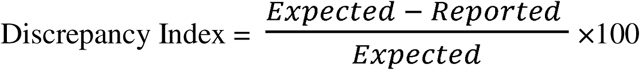

“Expected” was derived from LGA planning figures and ANC registration logs, while “Reported” represented final figures uploaded to Power BI.

### 2.6 Triangulated Interpretation

To contextualize numerical anomalies, qualitative data from STF field notes were thematically analyzed. Notes were categorized under key codes such as “tool deployment delay,” “supply stockouts,” “non-reporting,” and “logistics barrier.” These themes were then cross-referenced with LGA-level discrepancy scores to establish associations between operational issues and coverage outcomes. This mixed-method integration increased the interpretive depth and reliability of findings (Bhutta *et al.,* 2018; Gomes *et al.,* 2020).

### 2.7 Ethical Considerations

This study received ethical approval from; Bayelsa State Primary Health Care Board Research Ethics Committee (BSPHCBREC) Bayelsa State Primary Health Care Board, Yenagoa, Nigeria. Decision made; The BSPHCBREC granted full ethical approval (Approval No; Ref: BSPHCB/ERC/2025/112) for this study on 2nd of June 2025, waiving the need for individual participant consent as the research involved secondary analysis of anonymized programmatic data from the Maternal and Child Health Week (MCHW) OPS Room Report. All analysis was conducted in compliance with WHO guidelines on ethical use of health data and Nigeria’s National Health Research Ethics Code (WHO, 2020)

### 2.8 Limitations

The study is subject to several limitations. First, the reliance on secondary data implies that the analysis is constrained by the accuracy and completeness of routine service records. In several LGAs, STF reporting was incomplete, and MMS-specific entries were often merged with IFA data, limiting the precision of separate coverage estimates. Secondly, the lack of disaggregated data by maternal age, gestation stage, or ANC visit number restricted more granular analysis.

Finally, while STF notes were qualitatively analyzed, the absence of a formal sampling or structured interview guide for these narratives limited their depth and consistency.

## 3.0 RESULTS

This section presents the findings from the analysis of the June 2025 MNCH Week OPS Room Report in Bayelsa State, with particular attention to the coverage of maternal micronutrient supplementation (MMS), its comparison to other health interventions, and the systemic challenges that influenced its delivery and documentation. All data reflect validated entries compiled on the final day of the campaign period, as confirmed by the Power BI dashboard and field monitoring summaries.

### 3.1 Coverage of Maternal Micronutrient Supplementation (MMS) Among Pregnant Women

The available data indicate that a total of 2,197 pregnant women received either Iron-Folate or Multiple Micronutrient Supplements during the campaign. The estimated antenatal cohort for the state during the same period was 6,982. When evaluated against this estimate, coverage for maternal supplementation was considerably low. This suggests that only about one in three pregnant women benefitted from antenatal micronutrient supplementation during the June 2025 MNCH Week in Bayelsa.

The situation is particularly concerning given Bayelsa State’s high maternal anemia burden, with regional estimates indicating prevalence rates exceeding 60% among pregnant women, similar to national figures reported in the Nigeria Demographic and Health Survey (NDHS, 2018) and supported by Bayelsa State Health Surveillance Bulletins (NDHS, 2018; Bayelsa State Ministry of Health, 2022).

The incomplete recording and limited disaggregation between IFA and MMS interventions also meant that the true reach of the MMS strategy could not be fully verified. The summary of key indicators for maternal supplementation coverage is provided in Table 1 below.

This low level of coverage is further visualized in the figure below, which illustrates the proportion of pregnant women who received supplementation against those who did not. The distribution clearly shows a substantial implementation gap in reaching the full antenatal population with micronutrient interventions during the campaign.

**Figure 1:**
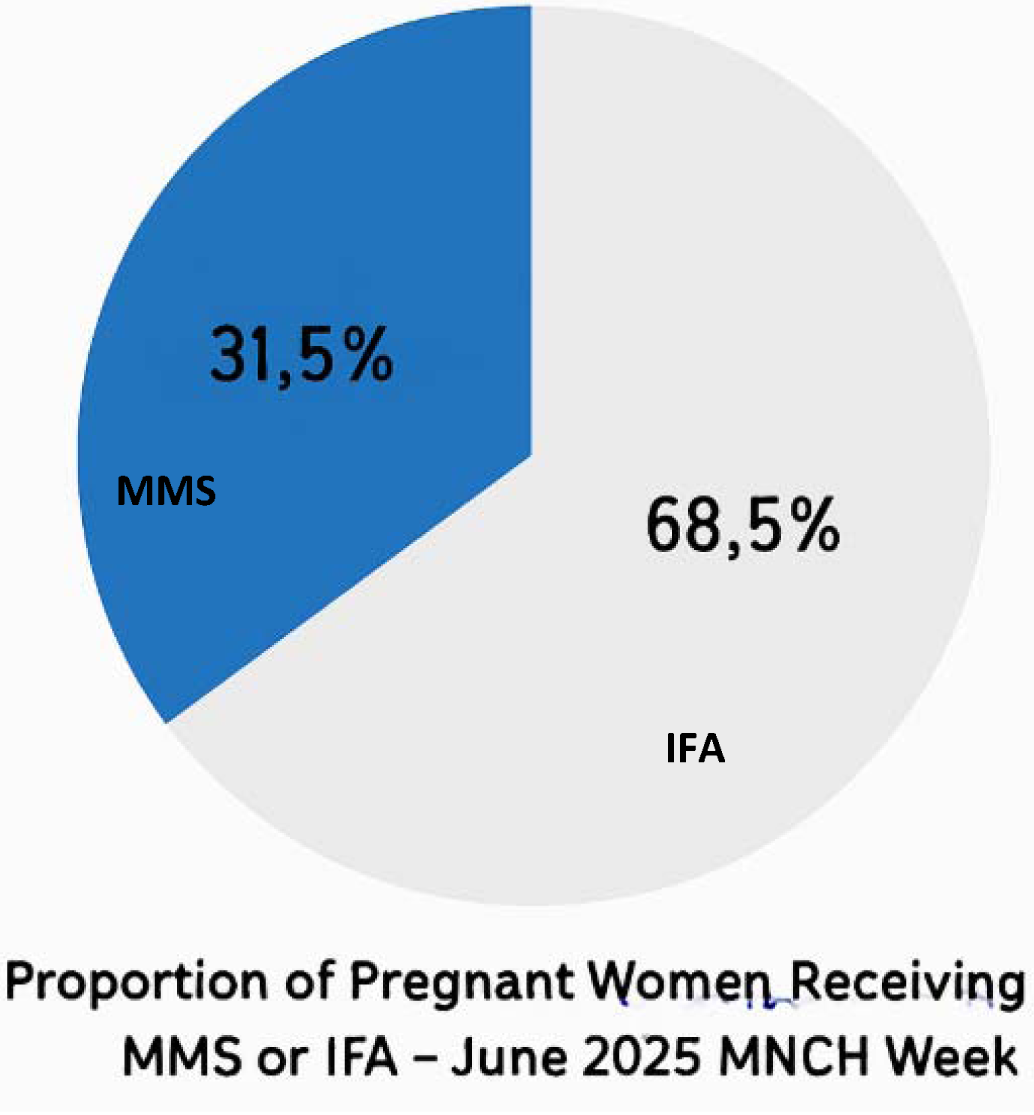
Proportion of Pregnant Women Receiving MMS or IFA – June 2025 MNCH Week (OPS Room Report, 2025)

### 3.2 Comparison with Other MNCH Interventions

To provide context for the performance of maternal supplementation during the campaign, its coverage was compared with other interventions delivered under the same MNCH Week platform. The analysis focused on Vitamin A supplementation and Deworming among children aged 6 to 59 months. The results show that Vitamin A supplementation achieved 83 percent coverage, with 279,996 children reached out of a target population of 337,215 (Oweibia *et al.,* 2025). Deworming achieved 55 percent coverage, with 185,197 children treated. Compared to these interventions, MMS performance was substantially lower. The maternal supplementation coverage of just over 31 percent highlights a major implementation shortfall and suggests either deprioritization, supply challenges, or weak integration into the broader campaign structure. The comparative results are summarized in Table 2.

**Table 2:**
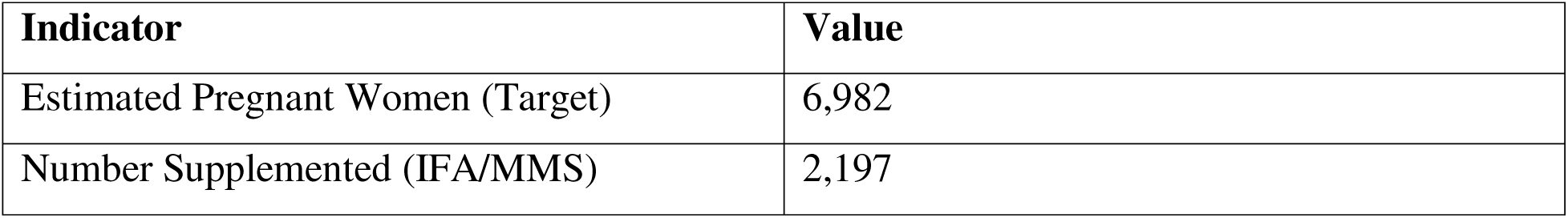
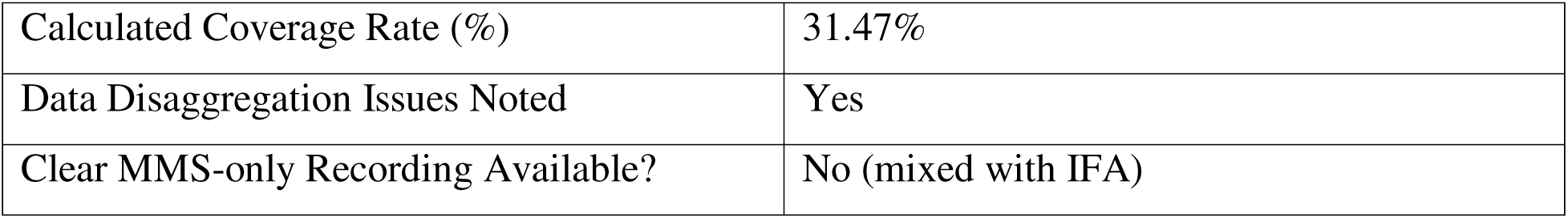
Maternal Micronutrient Supplementation Coverage – Bayelsa State (OPS Room Report, 2025)

The comparative difference is clearly depicted in the visual representation below. The disparity highlights not only the lower reach of maternal supplementation but also the potential systemic favoring of long-established child-focused interventions during campaign planning and logistics execution.

**Figure 2:**
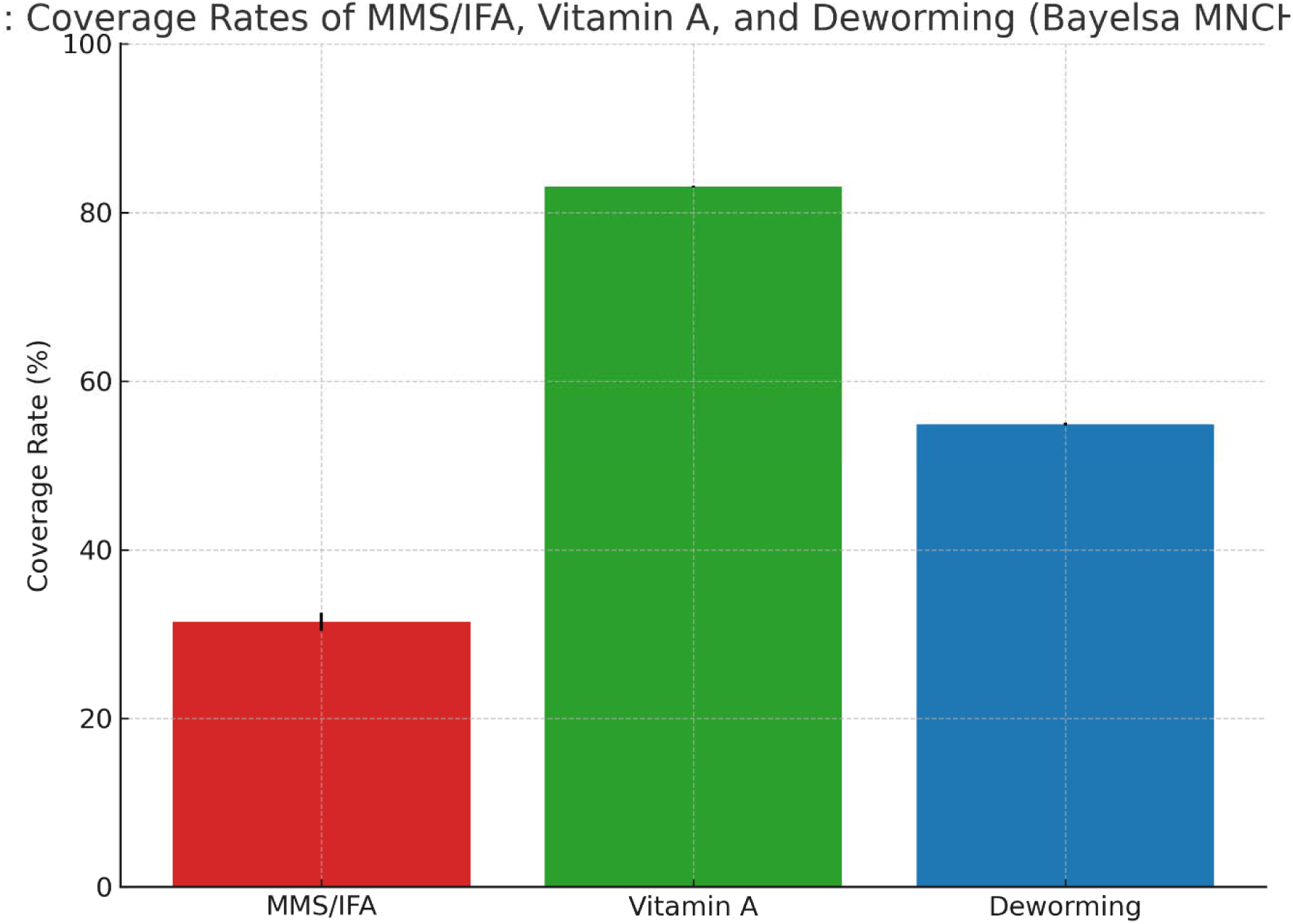
Coverage Comparison – MMS vs. Vitamin A and Deworming (OPS Room Report, 2025)

This contrast suggests a need for more equitable prioritization across maternal and child health services within the MNCH Week structure.

### 3.3 Operational and Systemic Challenges in MMS Delivery

Beyond quantitative underperformance, several systemic issues emerged from the OPS Room Report that help explain the low coverage figures observed. Field notes from State Technical Facilitators indicated that data collection tools for MMS were either not distributed before the start of the campaign or arrived late in multiple LGAs. As a result, many facilities lacked the standardized registers required to document MMS-specific data. In addition, campaign rollout was delayed in areas like Southern Ijaw LGA, where terrain-related challenges and logistic shortfalls disrupted timely initiation of services. STF teams in other LGAs, including Nembe and SILGA, failed to upload complete reports, further undermining data reliability and accountability.

In multiple instances, there was no clear distinction between Iron-Folate and MMS supplements in reporting formats, making it impossible to isolate and evaluate the performance of MMS as a distinct intervention.

These systemic documentation and delivery challenges are consistent with patterns observed in fragile health systems, where weak governance, poor data integration, and inadequate supply planning contribute to suboptimal service delivery (Kruk et al., 2018; WHO, 2020). These issues are summarized in Table 3.

**Table 3:** Comparative Coverage of MMS vs. Other Interventions (OPS Room Report, 2025)

| **Intervention** | **Target Population** | **Doses Administered** | **Coverage (%)** |
| --- | --- | --- | --- |
| MMS/IFA (Pregnant Women) | 6,982 | 2,197 | 31.5% |
| Vitamin A (Children) | 337,215 | 279,996 | 83.0% |
| Deworming (Children) | 337,215 | 185,197 | 55.0% |

**Table 4:** Documented Operational Challenges Affecting MMS Reporting (OPS Room Notes, 2025)

| Challenge Type | Description |
| --- | --- |
| Tool Deployment Delay | MMS registers not distributed before campaign start |
| STF Reporting Inconsistency | Missing entries from Nembe and SILGA |
| No Disaggregated Reporting | MMS and IFA data merged without distinction |
| Delayed Rollout | Southern Ijaw started campaign activities late |
| Logistic Gaps | Supply chain disruption in at least 3 LGAs |

The geographical distribution of reporting and rollout delays is illustrated in the figure below. These gaps were concentrated in hard-to-reach LGAs, especially those with riverine topography and limited infrastructure, which further constrained the delivery of ANC-linked interventions like MMS.

**Figure 3:**
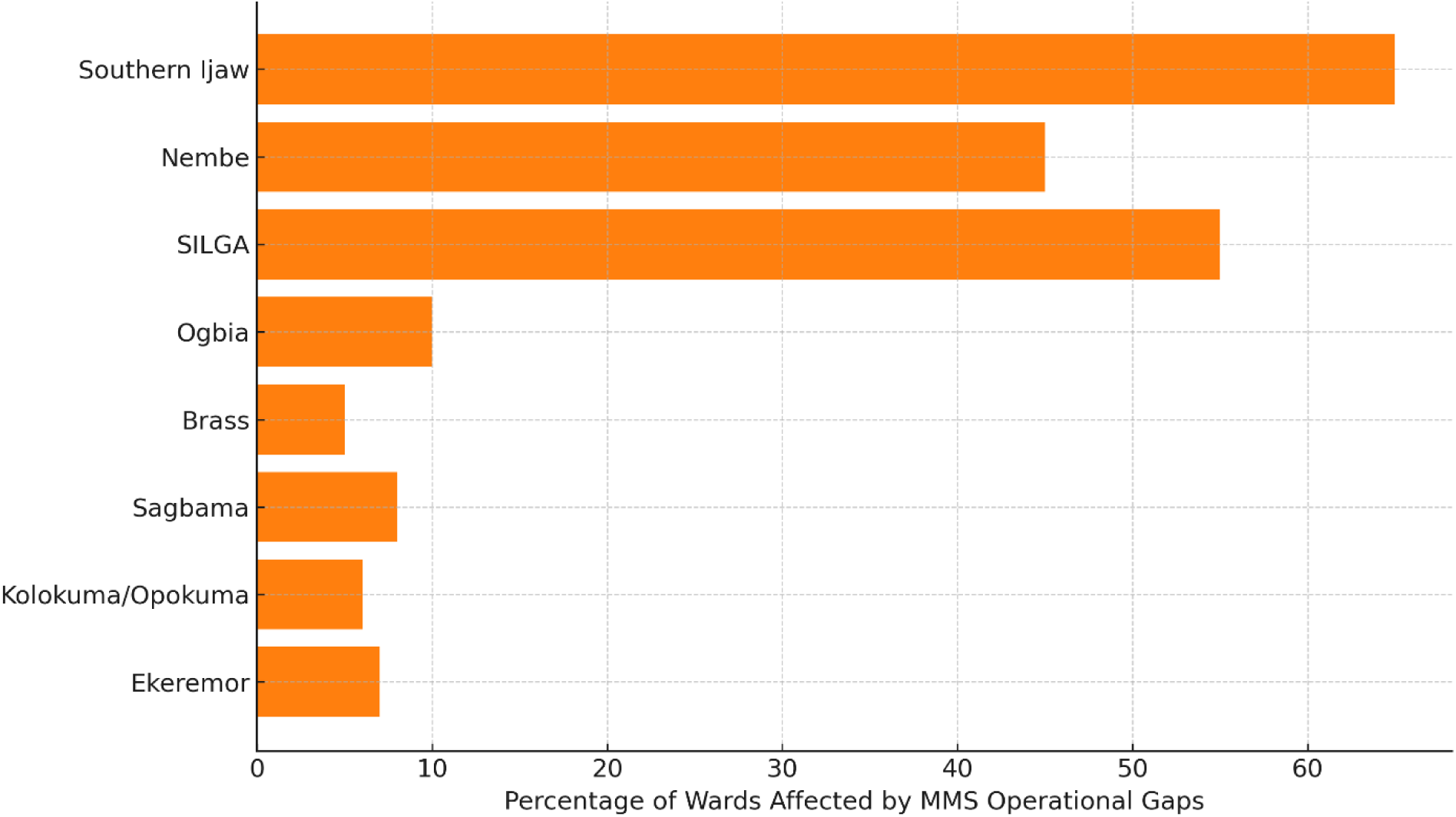
Bayelsa State LGAs with Known Reporting Gaps (OPS Room Report, 2025)

In summary, the results from the June 2025 MNCH Week reveal a dual-layer challenge in maternal micronutrient supplementation. On one hand, the quantitative data show low coverage and poor reach of pregnant women with MMS. On the other, operational evidence indicates that these figures likely reflect not just logistical failures but systemic weaknesses in documentation, planning, and integration of maternal services into campaign delivery.

## 4.0 DISCUSSION

### 4.1 Coverage of MMS/IFA: Systemic Causes and Interpretations

The maternal micronutrient supplementation (MMS/IFA) coverage rate of 31.5% observed during the June 2025 MNCH Week in Bayelsa State reveals critical gaps in the integration of maternal nutrition services into Nigeria’s campaign-based public health delivery structure. This rate, while reflective of some level of outreach success, falls significantly below optimal thresholds for population-level impact and underscores broader structural inefficiencies in service planning, documentation, and execution.

A major factor contributing to this suboptimal coverage is the systemic conflation of IFA and MMS in data collection and reporting. The lack of disaggregated registers led to the blending of figures, obscuring the true performance of MMS-specific services. This issue aligns with findings from other low- and middle-income countries (LMICs), where newly introduced maternal interventions often lack programmatic visibility due to inadequate tracking tools and weak integration at the point of care (Bhutta *et al.,* 2018; Gomes *et al.,* 2020).

Health worker familiarity and comfort with IFA further contributed to the underutilization of MMS. Studies have shown that providers often default to known practices in the absence of retraining or updated clinical protocols. Krause et al. (2021) found that inconsistent policy communication and insufficient training were major barriers to MMS uptake in ANC services across West Africa. In Nigeria, IFA is well-established in essential drug lists, while MMS remains relatively new and is still awaiting full inclusion in standard ANC guidelines (FMOH, 2023).

Beneficiary awareness and community demand for MMS also remain low. In many underserved communities, pregnant women are not informed about MMS or its benefits, often perceiving any antenatal supplement as generic vitamins. This mirrors findings from the 2018 Nigeria Demographic and Health Survey (NDHS) qualitative reports, which indicated poor differentiation between supplements among ANC clients (National Population Commission & ICF, 2019). Awareness strategies tailored to the local context are essential for uptake. Evidence from Nepal and Cambodia shows that integrating Social and Behavior Change Communication (SBCC) into routine ANC significantly increased MMS adherence (Nguyen *et al.,* 2019)

Ultimately, the results indicate that programmatic innovation alone, such as introducing MMS, is insufficient unless backed by systemic readiness. As emphasized in the Consolidated Framework for Implementation Research (CFIR), intervention success depends not only on efficacy, but also on contextual fit, implementation fidelity, and health system readiness (Damschroder *et al.,* 2009). Without appropriate tools, training, feedback loops, and supervision structures, even globally recommended interventions will fail to achieve impact.

### 4.2 Comparative Service Prioritization: Maternal vs. Child Interventions

The sharp contrast between MMS/IFA coverage (31.5%) and the much higher coverage of Vitamin A (83%) and Deworming (55%) during the same campaign week illustrates a longstanding imbalance in the prioritization of child versus maternal interventions. This discrepancy goes beyond logistics, it reflects decades of historical focus and global investment skewed toward child survival initiatives.

Vitamin A supplementation has benefitted from a well-documented history of global investment by WHO, UNICEF, and GAVI, with over 20 years of dedicated funding, robust supply chains, and widespread community mobilization (WHO, 2020; UNICEF, 2022). The ease of delivery, oral drops requiring minimal training, has made it a staple in child health campaigns and often bundled with immunization outreach.

In contrast, maternal interventions, particularly MMS, require more clinical judgment and programmatic attention. Proper MMS delivery involves dosage explanation, side effect counseling, and ANC-linked follow-up, all of which slow campaign workflows and may be seen as obstacles in fast-paced outreach programs (Young *et al.,* 2022). Without dedicated protocols and staff incentives, maternal services tend to be overlooked or absorbed into more generic reporting categories, as the Bayelsa case demonstrates.

Global comparisons reinforce this finding. In Vietnam, national MMS coverage only exceeded 80% after the Ministry of Health restructured ANC services to include new digital tracking tools, community health worker incentives, and dedicated government funding (Rana & Clift, 2023). In Bangladesh, leveraging a decentralized health system and real-time dashboards boosted maternal supplement coverage to over 70% in rural regions (Ahmed *et al.,* 2021).

These examples affirm that maternal health cannot be an afterthought to child-focused efforts. It requires independent planning, budget lines, monitoring structures, and public health visibility. Nigeria’s maternal health programming must evolve from episodic inclusion within child health events to full-fledged vertical integration with its own service delivery framework (Oweibia *et al.,* 2025).

### 4.3 Operational Barriers and Regional Inequities in Bayelsa

The challenges observed in LGAs such as Southern Ijaw, Nembe, and SILGA reflect a broader national dilemma: health inequities driven by geographical inaccessibility. In Bayelsa’s riverine terrain, most communities can only be reached by boat, making supply delivery, staff deployment, and data reporting exceptionally difficult. These patterns of exclusion are not new studies have long shown that terrain-related isolation in Nigeria exacerbates maternal and child health disparities (FMOH, 2022; UNFPA, 2022).

To overcome such barriers, other countries have piloted innovative logistics solutions. In Rwanda and Ghana, drone-supported deliveries of vaccines and medical commodities have dramatically improved reach to remote communities (Zipline, 2021; Gavi, 2022). Similarly, Sierra Leone’s mobile boat clinics have proven successful in delivering maternal and newborn health services across swampy and island terrains (PATH, 2021). Nigeria’s riverine states, particularly those in the Niger Delta, can benefit from adapting these models.

Another major issue identified in Bayelsa was the delayed deployment of MMS-specific tools. Facilities lacked proper registers or digital forms to capture intervention-specific data, leading to merged reporting under IFA. Without granular documentation, coverage data lose reliability and cannot inform planning. As emphasized by WHO’s Health Data Quality Toolkit, accurate tool deployment and feedback loops are non-negotiable for campaign integrity (WHO, 2017).

The discussion would be incomplete without addressing gender dynamics, which shape maternal service uptake. In many Nigerian communities, health decisions for pregnant women are influenced or controlled by spouses or older family members, limiting women’s autonomy. Research shows that male involvement in maternal health increases ANC attendance and supplement adherence (UNFPA, 2021; Olorunsaiye *et al.,* 2020). Community-based strategies that engage men as allies through religious, social, and workplace channels can significantly shift household decision-making and increase maternal health access.

In conclusion, the Bayelsa experience illustrates a multi-layered challenge: a health system strained by geography, outdated processes, weak accountability, and sociocultural inertia. If Nigeria is to reach its SDG 2 and 3 targets ending malnutrition and reducing maternal mortality then systemic reforms, context-responsive delivery models, and gender-inclusive health promotion must become core components of national maternal health strategies.

## 5.0 CONCLUSION AND RECOMMENDATIONS

### 5.1 Conclusion

This study assessed the coverage and implementation bottlenecks of maternal micronutrient supplementation (MMS) during the June 2025 Maternal and Child Health (MNCH) Week in Bayelsa State. Despite a well-established campaign platform, coverage for MMS/IFA among pregnant women was only 31.5%, a figure that falls far below both national and global targets. In contrast, Vitamin A and Deworming services reached 83% and 55%, respectively, highlighting a performance disparity across service categories.

The findings underscore systemic deficiencies in Nigeria’s maternal nutrition programming, including the absence of disaggregated data collection tools, delayed rollout, and logistical constraints, especially in geographically isolated local government areas (LGAs) such as Southern Ijaw, Nembe, and SILGA. These constraints mirror broader challenges facing LMICs, where maternal health interventions often receive less operational support compared to child-focused services (WHO, 2020; UNICEF, 2022).

While Nigeria has demonstrated strong momentum in scaling child health interventions, maternal nutrition continues to be deprioritized within both routine and campaign service delivery models. The episodic nature of campaign-based MMS delivery, without parallel investments in tools, training, supply chains, and community demand generation, limits programmatic effectiveness. Furthermore, subnational inequities and sociocultural barriers persist as major determinants of maternal health access (FMOH, 2023; UNFPA, 2021).

To drive progress, MMS must be repositioned as a core component of routine antenatal care (ANC), embedded within Nigeria’s essential maternal health package. Achieving coverage scale, equity, and sustainability will depend on cross-cutting reforms in digital monitoring, logistics innovation, and gender-responsive community engagement. Without these, Nigeria risks stalling progress toward national and global nutrition and health targets, including Sustainable Development Goals 2 and 3 (SDG Tracker, 2023; World Bank, 2022).

### 5.2 Recommendations

To ensure sustained improvement in maternal micronutrient supplementation (MMS) delivery in Bayelsa State, MMS should be institutionalized within routine antenatal care (ANC) services across all Primary Health Care (PHC) facilities, transitioning from campaign-based models to continuous service delivery. This aligns with the World Health Organization (WHO, 2020) and Global Nutrition Report (2021) recommendations on mainstreaming maternal nutrition. Routine integration will require the deployment of real-time, mobile-based dashboards and digital e-registers to capture disaggregated data on iron-folic acid (IFA) and MMS usage. Tools like these have proven effective in improving data quality and program responsiveness in other low- and middle-income countries (Ahmed *et al.,* 2021). Standardized, MMS-specific registers should also be designed and disseminated nationwide, distinct from traditional IFA templates, to enable better tracking of coverage metrics (FMOH, 2023; Gavi, 2022). Moreover, digital health tools should support monitoring maternal health trends and disparities during campaigns, as highlighted by Oweibia et al. (2025), who reported varying degrees of Vitamin A and deworming coverage during MNCH Weeks. Strengthening logistics through terrain-specific strategies like boat clinics, drone delivery, and pre-stocked kits, especially in hard-to-reach riverine areas of Bayelsa, is essential. These innovations have been successful in similar contexts across Ghana, Rwanda, and Sierra Leone (UNICEF, 2022; PATH, 2021).

Capacity building of frontline workers is another critical pillar. Pre-campaign trainings on MMS eligibility, dosing, and accurate digital data entry should be institutionalized. Such training has shown positive outcomes in WHO-supported trials (WHO, 2022) and can also address observed disparities in malnutrition screening across LGAs as reported by Oweibia et al. (2025d). Furthermore, community mobilization must adopt gender-sensitive and inclusive approaches that involve Ward Development Committees, male influencers, and faith-based organizations to drive uptake. Evidence from the NDHS (2018) and UNFPA (2021) underscores the role of male involvement in improving maternal health-seeking behaviors. To bolster accountability and data integrity, Local Government Area (LGA)-based monitoring teams should be deployed to conduct real-time audits and validate reported entries (PATH, 2021; FMOH, 2023). Coverage dashboards should be used to identify and target underperforming LGAs with additional resources and supervision (Gomes *et al.,* 2020). Structured after-action reviews involving State Task Forces (STFs), PHC managers, and community representatives should be conducted post-MNCH Weeks to foster adaptive learning (WHO, 2020). Finally, securing stable funding through dedicated MMS budget lines in state and national maternal health frameworks, including the Basic Health Care Provision Fund (BHCPF), is imperative for scaling up MMS efforts beyond pilot stages (Global Nutrition Report, 2021; World Bank, 2022). These recommendations align with broader maternal health policy frameworks and reflect insights from recent analyses of maternal health system performance and health intervention coverage in Nigeria (Oweibia *et al.,* 2025; Oweibia *et al.,* 2025).

## Data Availability

All data produced in the present work are contained in the manuscript

